# An 8-Week Oral Therapy with Ashwagandha (*Withania Somnifera*) Root Extract (600 mg/day) Improves Frailty and Quality of Life in Elderly: A Prospective, Randomized, Placebo-Controlled, Efficacy and Safety Study

**DOI:** 10.1101/2024.10.15.24315515

**Authors:** K. Sunil Naik, Gudla Muralidhar, Bade Srinivas Rao, Kalpana Wankhade

## Abstract

Frailty is a state of increased vulnerability resulting from aging due to cumulative decline in physiological system over a lifespan. Few pharmacological agents have been investigated for the treatment of frailty. In Ayurveda, Ashwagandha (*Withania Somnifera*) is a popular botanical medicine used for improvement in physical strength and mental stress and has a potential for treatment of frailty. This placebo-controlled study assessed the efficacy and safety of a capsule containing 300mg of Ashwagandha Root Extract (ARE) administered twice daily orally for 8 weeks. Fifty elderly subjects with a frailty score ≥7 based on Frailty Assessment and Screening Tool (FAST) were randomized in a 1:1 ratio to receive either Ashwagandha (ARE, n=25) or placebo (PL, n=25). Improvement in frailty after 8 weeks was assessed by FAST, 6-minute Walk Test (6MWT), Pittsburgh Sleep Quality Index (PSQI), Mini-Mental State Examination (MMSE) and Short Form Survey (SF-12) scores. Blood samples were collected at baseline and week 8 for estimation of C-Reactive Protein (CRP), cortisol and Creatinine Kinase (CK). Significant improvement (p<0.01) in scores for FAST, 6MWTs, PSQI, MMSE, and SF-12 were seen with ARE after 8 weeks. Also, significant improvements (p<0.05) were observed in CK and in CRP with ARE. These improvements were greater (p<0.05) with ARE as compared to placebo. ARE was well tolerated with no adverse effects, and no changes in hepatic and renal parameters. Thus, Ashwagandha root extract can be a valuable therapeutic option for improvement of health condition in frailty.

## INTRODUCTION

Aging and senescence are natural and unavoidable conditions in humans, and frailty is a rapidly increasing major health challenge in aging society. Geriatric frailty is found in 20-30% of the people over 75 years and it increases with aging.[1] The prevalence of frailty in India ranges from 11-58%.[2], [3], [4] Globally, due to increase in elderly population, management of aging and its associated frailty is a challenge for the healthcare systems.[5] Good health remains an unaccomplished goal in elderly, and frail individuals have a two-fold increased risk of complications compared to non-frail individuals.[6] Decline in physical strength in elderly has been attributed to the loss of skeletal muscle mass and muscle quality referred to as sarcopenia, resulting from anatomic and biochemical changes in the aging muscle.[7] There is a need to explore pharmacological and non-pharmacological interventions to prevent and improve frailty.

Alternate therapies such as Ayurvedic medicines including Rasayana herbs, Yoga, and exercises prove to be effective in the management of age-related health issues.[5] Ashwagandha (*Withania Somnifera*) commonly known as Indian winter berry or Indian Ginseng, is one of the most important traditional Ayurvedic medicine used in India since past 6000 years. It is an adaptogen and used for its wide range of health benefits to promote youthful state of physical and mental health. The root of Ashwagandha is regarded as tonic, aphrodisiac, narcotic, diuretic, anthelmintic, astringent, thermogenic and stimulant. It is commonly used in therapy for emaciation in children (when given with milk, it is the best tonic for children), debility in elderly, rheumatism, leucoderma, constipation, insomnia, nervous breakdown, goiter etc.[8] Pharmacologically active constituents of Ashwagandha root extract contain steroidal lactones, named Withanolides. Withania, a group of alkaloids isolated from the roots of plants, forms 38% of total weight of alkaloids as well as steroids, saponins, phenolics flavonoids, Phyto-phenols, and glycosides. It is widely used in traditional medicine formulations as an antipyretic, analgesic, adaptogenic, and anti-inflammatory agent.[9] It also has numerous health benefits, enhanced brain and nervous system function, improved memory, and promotion of a healthy sexual and reproductive balance. [10], [11] Ashwagandha is used as a general tonic and as an adaptogen to help the body cope with the stress and may be useful in Frailty. Ashwagandha root extract (ARE) can be a potential therapeutic option for improvement of frailty and quality of life in older adults. This placebo-controlled study evaluated the efficacy and safety of Ashwagandha root extract in elderly frail individuals.

## MATERIAL AND METHODS

### Study Design and Setting

This eight-week treatment period, prospective, randomized, double-blind, two-arm, placebo-controlled, parallel, comparative study evaluated the efficacy and safety of ARE in improving frailty and quality of life (QoL) in elderly men and women. The clinical study protocol and related documents were reviewed and approved by the Institutional Ethical Committee of Government Medical College and Government General Hospital, Srikakulam-532001, Andhra Pradesh, India. (Date of approval: January 31, 2022). The study was conducted in compliance with the good clinical practice guidelines (ICH GCP), New Drugs and Clinical Trials Rules 2019 (India), and Declaration of Helsinki (Taipei 2016). Approval was obtained from the institutional ethics committee prior to study procedures, and written informed consent was obtained from all the subjects prior to any study related procedures. The study was registered at clinical trials registry of India (CTRI/2022/02/040554; dt. 23 Feb. 2022). Reporting of the results of this study adheres to the Consolidated Standards of Reporting of Randomized Trials (CONSORT) criteria.

### Study participants

Elderly men and women aged 65 years or more with a baseline score of 7 or more on the Frailty Assessment and Screening Tool (FAST) scale, and willing to comply with the study activities were included. Those with known cognitive impairment having a Mini-Mental State Examination (MMSE) score of ≤20, those with known mental disorders, or receiving palliative treatment were excluded. Those with a history of alcohol or substance abuse, presence of any clinically significant cardiovascular, renal, metabolic, hematological, neurological, systemic, or other infectious disease or malignant tumor, or any other acute illness were also excluded. Those with a history of surgery within one year, consumption of any other dietary or nutritional supplements, multivitamins at least one month before the study, and history of allergy to any of the components of investigational product were an exclusion.

### Randomization and Blinding

Enrolled subjects were assigned a unique serial number and randomized (1:1 ratio) to receive a capsule containing either ARE (n=25) or placebo (PL, n=25). Enrolled participants received pre-sealed study medication packs based on their serial number. The study products (active and placebo) were identical and were of the same form and color. Being a double-blind study, sequentially numbered opaque sealed envelopes (SNOSE) were used to maintain blinding. It was ensured that the subjects as well as the researcher and the staff were blinded to the two treatments. Blinded randomization codes were maintained at the custody of investigator. No member of the study site (except principal investigator) nor the study monitor had access to the randomization code until the end of the study. Investigator could only break the code, in case of emergency. A non-affiliated researcher who was deprived of sight gathered the results of the assessments at each planned appointment.

### Interventions

Participants in the ARE group received capsule containing 300 mg of ARE (KSM-66 Ashwagandha root extract, Ixoreal Biomed, CA, US) orally twice daily after breakfast and dinner, with a glass of water, for a duration of eight weeks. The placebo (PL) group was administered an identical capsule containing 300 mg starch.

### Study outcomes

The primary outcome was a change from baseline in the Frailty Assessment and Screening Tool (FAST) scores at week 8. Secondary outcomes included change in the values for 6-minute walk test (6MWT), PSQI (Pittsburgh sleep quality index), MMSE (Mini-mental state examination), and SF-12 (Short-Form 12). Safety was assessed based on clinical adverse events, serum C-reactive protein (CRP), cortisol, creatinine kinase levels assessed at baseline and week 8.

### Study assessments

The study comprised of two on-site visits (baseline visit - day 1, end of study visit - week 8) and one telephonic visit (week 4). A telephonic follow-up visit was done to ensure adherence to drug therapy and protocol, as well as to gather safety data regarding adverse events (AEs).

#### Frailty Assessment and Screening Tool (FAST)

FAST is a validated tool with good psychometric properties commonly used to assess frailty in older people.[12] It consists of a total of 14 items pertaining to 10 domains. Time required for completion of this scale by the clinician is less than 15 minutes. The highest total score is up to 14, and higher score denotes a frailer condition.

#### 6-minute walk test (6MWT)

The 6-minute walk test is a sub-maximal exercise test used to assess aerobic capacity and endurance.[13] The distance covered over a time of 6 minutes is used as the outcome by which changes in performance capacity are compared. The test evaluates the functional capacity of the individual and it provides valuable information regarding all the systems during physical activity, including pulmonary and cardiovascular systems, blood circulation, neuromuscular units, body metabolism, and peripheral circulation. The objective of this test is to walk as far as possible for 6 minutes. Six minutes is a long time to walk, so patients exert themselves and probably get out of breath or become exhausted.

#### Pittsburgh Sleep Quality Index (PSQI)

PSQI is an effective instrument used to measure the quality and patterns of sleep-in adults.[14] It differentiates “poor” from “good” sleep quality by measuring seven areas (components): subjective sleep quality, sleep latency, sleep duration, habitual sleep efficiency, sleep disturbances, use of sleeping medications, and daytime dysfunction over the last month. The questionnaire contains 19 self-related questions that form 7 “components” scores, each of which has 0-3 points. There are 5 questions rated by the sleeping partner or a roommate. Only self-related questions are included in scoring and rated on 0–3-point scale. (0 indicates no difficulty and 3 indicates severe difficulty). Single global score is obtained by adding 7-component (0-21-point scale) score (0 indicates no difficulty and 3 indicates severe difficulty in areas).

#### Mini-Mental State Exam (MMSE)

The MMSE is a widely used test of cognitive function among the elderly; it includes tests of orientation, attention, memory, language, and visual-spatial skills. It can be used to indicate the presence of cognitive impairment, such as in a person with suspected dementia or following a head injury. The examination has been validated in several populations. Scores of 25-30 out of 30 are considered normal; the National Institute for Health and Care Excellence (NICE) classifies 21-24 as mild, 10-20 as moderate and <10 as severe impairment.[15], [16]

#### Short Form Survey (SF-12) scale for Quality of Life (QoL)

It is used to assess quality of life for the assessment of impact of health on an individual’s everyday life. It consists of 12-question survey using 8 domains such as limitations in physical activities due to health problem, in social activities due to physical and emotional problems, in usual role activities due to bodily pain, vitality (energy and fatigue) and general health perceptions. It is assessed by scores ranging from 0-100 with higher scores indicating better physical and mental health.[17]

#### Laboratory assessments

Effects of ARE on blood, liver and kidney were assessed by laboratory assessments at baseline and week 8. Blood samples were evaluated for C-reactive protein (CRP), cortisol, and creatinine kinase levels. Hematology (cell counts, hemoglobin, and hematocrit), and serum biochemistry was done. Hepatic parameters included serum bilirubin, total cholesterol, and alkaline phosphatase, whereas renal parameters included blood urea nitrogen (BUN), creatinine, aspartate transaminase (AST), and alanine transaminases (ALT).

### Safety assessments

Adverse events (AE) reported by the patients and children were recorded based on their recall during telephonic follow-up and site visits. Treatment-emergent AEs were described by counts and percentages, and summaries were presented both by treatment and overall. Results from physical examinations were listed, and vital signs results were summarized using descriptive statistics. A telephonic follow-up visit (day 28) was done to ensure adherence to drug therapy and protocol, as well as to gather safety data regarding adverse events (AEs).

### Sample size

The sample size was calculated based on the meaningful improvements in gait speed (estimated at 0.05 to 0.2 m/s) predicting positive outcomes on a population level. An improvement of 0.05 to 0.2 m/s in the gait speed is considered clinically significant. The sample size was estimated based on hypothesis that the intervention group had a 0.05 m/s improvement from baseline in the gait speed. A sample size of 21 in each arm (total 42 patients) provides 90% power to detect this change. Expecting a drop-out of 15% patients’ post-randomization, a total sample size was set at 50 (25 patients in each arm).

### Statistical methods and data analysis

Since all patients completed the study, data of all patients was included for analysis of efficacy and safety outcomes. For continuous variables, the number of subjects, mean, median, standard deviation (SD), and minimum and maximum values were presented. Paired t-test was used for the within-group (baseline – week 8) comparison, a two-independent samples t-test was used for the between-group (ARE vs. Placebo) comparison at baseline and week 8 and ANCOVA was used for the comparison of between-group (ARE vs. Placebo) after controlling for baseline. All relevant statistical calculations were completed using SPSS software (Version 21, IBM Corporation, USA). The obtained analysis outcomes for ranking data and scores were tabulated as mean ± SD. To ensure adherence to best statistical practices, 95% Confidence Intervals (CI) were utilized in the study. All analyses were done using two-sided tests and a p-value of less than 0.05 was considered the threshold to claim statistical significance. For categorical variables, the number and percentage of subjects within each category (with category for missing data as needed) of the parameter were presented and P-values were determined by Chi-square test between the groups (ARE vs. Placebo).

## RESULTS

A total of 50 participants (31 males and 19 females) were allocated equally into two groups (figure-1). All participants completed the study as per the protocol and were included in the ITT/PP datasets for efficacy and safety analysis.

**Figure 1.**
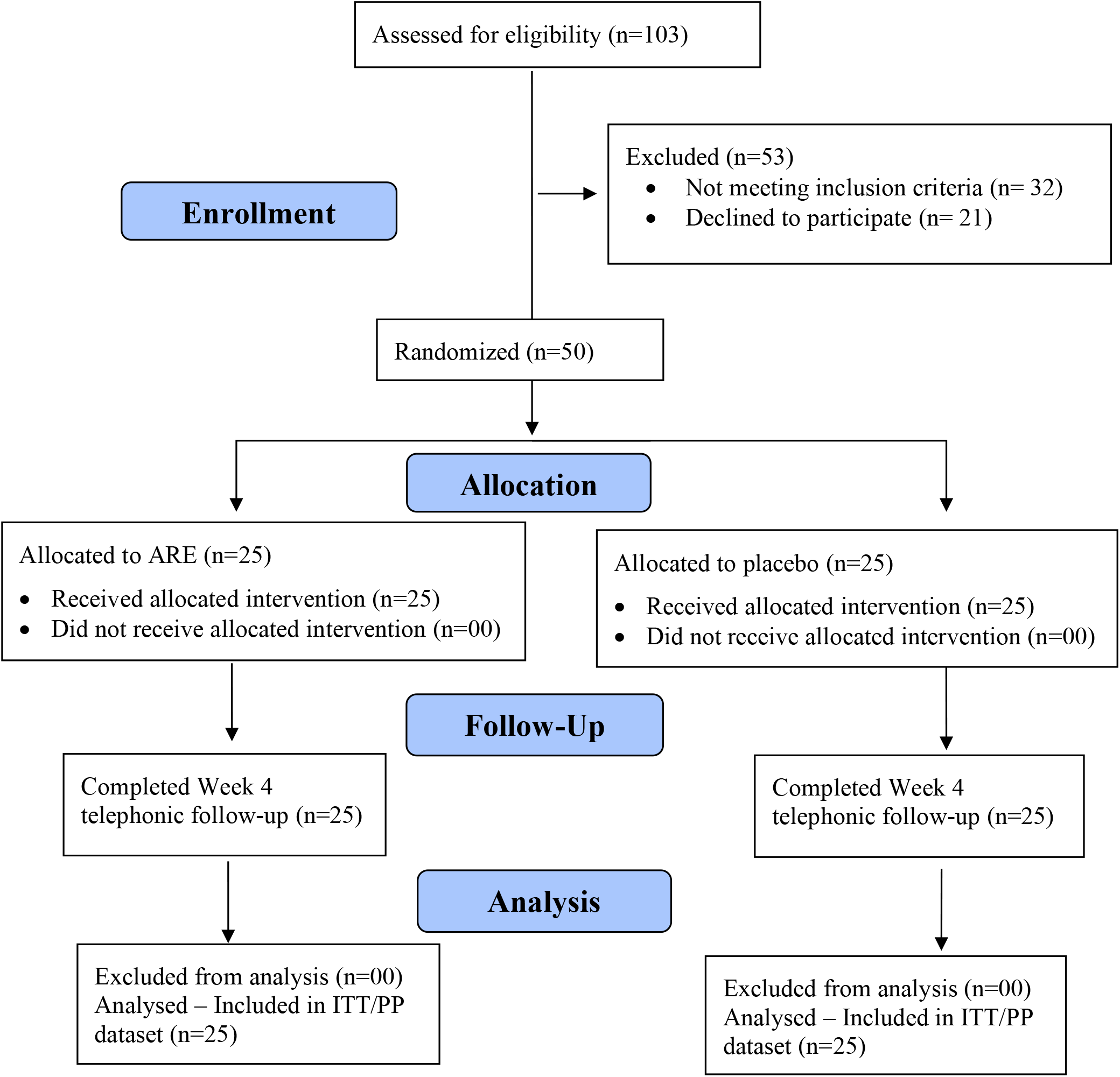
CONSORT 2010 Flow Diagram

**Figure 2.**
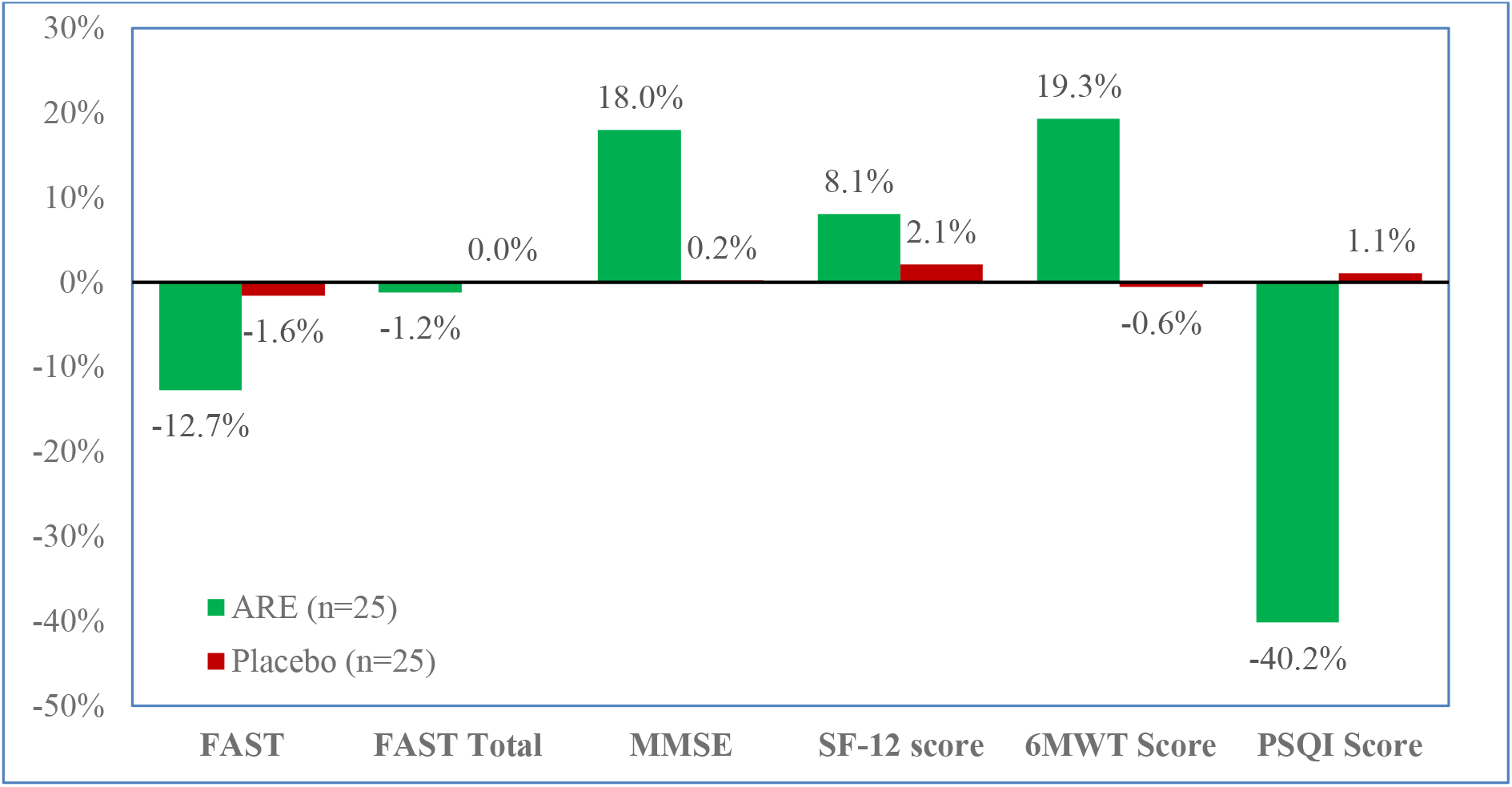
Percent change from baseline after 8 week therapy in two groups

### Profile of patients and baseline data in ITT dataset

The two groups in the ITT dataset (n=50) were comparable (p>0.05) with respect to the profile of participants (table-1), and baseline data for different laboratory parameters (table-3).

**Table 1.** Demography at baseline in ITT dataset (n=50)

|  | ARE (n=25)<br>Mean (SD) | Placebo (n=25)<br>Mean (SD) | *t* | *p* |
| --- | --- | --- | --- | --- |
| **Demography** |  |  |  |  |
| Age (yrs.) | 64.3 (1.1) | 64.0 (0.8) | 0.953 | 0.346 |
| BMI (kg/m<sup>2</sup>) | 22.9 (2.4) | 24.0 (2.1) | -1.643 | 0.107 |
| | No. (%) | No. (%) | $\chi^2$ | *p* |
| **Sex** |  |  |  |  |
| Male | 15 (60.0%) | 16 (64.0%) | 0.085 | 0.771 |
| Female | 10 (40.0%) | 9 (36.0%) |  |  |
| **Marital Status** |  |  |  |  |
| Married | 23 (92.0%) | 19 (76.0%) | 2.381 | 0.123 |
| Widow | 2 (8.0%) | 6 (24.0%) |  |  |
| **Education** | 0.00% |  |  |  |
| SSC | 3 (12.0%) | 1 (4.0%) | 3.41 | 0.182 |
| HSC | 4 (16.0%) | 1 (4.0%) |  |  |
| Degree and above | 18 (72.0%) | 23 (92.0%) |  |  |
| **Nutritional Status** |  |  |  |  |
| Malnutrition | 3 (12.0%) | 0 (0.0%) | 3.243 | 0.198 |
| Undernutrition | 3 (12.0%) | 4 (16.0%) |  |  |
| Overnutrition | 19 (76.0%) | 21 (84.0%) |  |  |
| **Occupation** |  |  |  |  |
| Office or business | 19 (76.0%) | 17 (68.0%) | 0.535 | 0.765 |
| Skilled or semi-skilled | 5 (20.0%) | 6 (24.0%) |  |  |
| Unemployed | 1 (4.0%) | 2 (8.0%) |  |  |
| **Cohabitation** |  |  |  |  |
| Alone | 2 (8.0%) | 3 (12.0%) | 0.892 | 0.640 |
| Living with spouse | 4 (16.0%) | 2 (8.0%) |  |  |
| With spouse and children | 19 (76.0%) | 20 (80.0%) |  |  |
| **Income (INR per annum)** |  |  |  |  |
| <5 lakhs | 21 (84.0%) | 24 (96.0%) | 1.960 | 0.157 |
| 5 to 15 lakhs | 4 (16.0%) | 1 (4.0%) |  |  |

ARE: Ashwagandha root extract; BMI: Body mass index; HSC: Higher secondary school certification; INR: Indian rupee; ITT: Intent-to-treat; SSC: Secondary school certification

### FAST, 6MWT, MMSE, PQSI and SF-12 scores

The two groups were similar (p>0.05) with respect to the baseline values for FAST score, 6MWT, MMSE total score, PSQI score, and SF-12 score (table-2). The FAST scores were lower with ARE at week 8 (p=0.004), and greater reduction (p=0.020) was seen with ARE compared to placebo. Thus, significant reduction in FAST score was seen with ARE (p=0.008) but not with placebo (p>0.05) from baseline to week 8, which indicates greater improvement in frailty with ARE. Similarly, significant improvements in 6MWT, MMSE, PSQI and SF-12 scores were seen with ARE (p<0.0001) at the end of week 8, whereas no improvements were seen with placebo (p>0.05). Between group comparisons show better improvements (large effect sizes) for 6MWT score (p<0.0001), MMSE score (p<0.0001), PSQI score (p<0.0001) and SF-12 (p<0.0001) score with ARE compared to placebo (table-3). Greater percent improvement from baseline in different parameters was observed after 8 week therapy with ARE as compared to placebo (figure-1).

**Table 2.** Comparison between groups for different parameters.

| Parameter |  | ARE (n=25)<br>*Mean (SD)* | Placebo (n=25)<br>*Mean (SD)* | Difference between groups<br>*Mean (95% C.I.)* | Unpaired t-test<br>*t (p)* | Effect size<br>*Cohen's d* |
| --- | --- | --- | --- | --- | --- | --- |
| **Assessment scales** |  |  |  |  |  |  |
| FAST score | *Baseline* | 7.56 (1.19) | 7.60 (0.76) | -0.04 (-0.61 to 0.53) | -0.141 (0.888) | -0.040 |
|  | *Week 8* | 6.60 (1.29) | 7.48 (0.65) | -0.88 (-1.46 to -0.30) | -3.041 (0.004) | -0.860 |
|  | *Change in wk. 8* | -0.96 (1.67)* | -0.12 (0.53)<sup>#</sup> | -0.84 (-1.54 to -0.14) | -2.398 (0.020) | -0.678 |
| 6MWT score | *Baseline* | 119.08 (17.75) | 123.04 (16.99) | -3.96 (-13.84 to 5.92) | -0.806 (0.424) | -0.228 |
|  | *Week 8* | 142.08 (32.47) | 122.32 (15.89) | 19.76 (5.22 to 34.30) | 2.733 (0.009) | 0.773 |
|  | *Change in wk. 8* | 23.00 (25.84)** | -0.72 (13.65)<sup>#</sup> | 23.72 (11.97 to 35.47) | 4.059 (<0.001) | 1.148 |
| MMSE score | *Baseline* | 20.68 (0.90) | 20.48 (0.77) | 0.20 (-0.28 to 0.68) | 0.844 (0.403) | 0.239 |
|  | *Week 8* | 24.40 (4.43) | 20.52 (1.23) | 3.88 (2.03 to 5.73) | 4.216 (<0.001) | 1.192 |
|  | *Change in wk. 8* | 3.72 (4.41)** | 0.04 (0.93)<sup>#</sup> | 3.68 (1.87 to 5.49) | 4.081 (<0.001) | 1.154 |
| PSQI score | *Baseline* | 15.44 (1.92) | 14.84 (2.15) | 0.60 (-0.56 to 1.76) | 1.040 (0.303) | 0.294 |
|  | *Week 8* | 9.24 (5.99) | 15.00 (2.04) | -5.76 (-8.31 to -3.21) | -4.548 (<0.001) | -1.286 |
|  | *Change in wk. 8* | -6.20 (5.96)** | 0.16 (1.21)<sup>#</sup> | -6.36 (-8.81 to -3.91) | -5.230 (<0.001) | -1.479 |
| SF-12 score | *Baseline* | 19.84 (2.21) | 19.28 (2.46) | 0.56 (-0.77 to 1.89) | 0.847 (0.401) | 0.240 |
|  | *Week 8* | 21.44 (1.23) | 19.68 (1.77) | 1.76 (0.89 to 2.63) | 4.081 (<0.001) | 1.154 |
|  | *Change in wk. 8* | 1.60 (1.83)** | 0.40 (1.08)<sup>#</sup> | 1.20 (0.35 to 2.05) | 2.828 (0.007) | 0.800 |
| **Specific laboratory parameters** |  |  |  |  |  |  |
| CRP | *Baseline* | 3.20 (1.08) | 3.31 (1.11) | -0.11 (-0.73 to 0.51) | -0.362 (0.719) | -0.102 |
|  | *Week 8* | 3.28 (1.03) | 3.46 (1.06) | -0.18 (-0.77 to 0.42) | -0.598 (0.553) | -0.169 |
|  | *Change in wk. 8* | 0.08 (1.32)** | 0.14 (1.55)<sup>#</sup> | -0.06 (-0.88 to 0.76) | -0.157 (0.876) | -0.044 |
| Serum Cortisol | *Baseline* | 209.64 (57.53) | 225.28 (75.71) | -18.20 (-54.38 to 17.98) | -1.011 (0.317) | -0.286 |
|  | *Week 8* | 206.64 (57.53) | 225.28 (75.71) | -18.64 (-56.88 to 19.60) | -0.980 (0.332) | -0.277 |
|  | *Change in wk. 8* | -18.04 (56.40)** | -2.56 (52.65)<sup>#</sup> | -15.48 (-46.51 to 15.55) | -1.003 (0.321) | -0.284 |
| CK | *Baseline* | 2.60 (1.35) | 2.28 (1.06) | 0.32 (-0.37 to 1.01) | 0.930 (0.357) | 0.263 |
|  | *Week 8* | 2.56 (1.00) | 2.36 (1.08) | 0.20 (-0.39 to 0.79) | 0.680 (0.500) | 0.192 |
|  | *Change in wk. 8* | -0.04 (1.54)** | 0.08 (1.47)<sup>#</sup> | -0.12 (-0.98 to 0.74) | -0.282 (0.779) | -0.080 |

\* $p < 0.01$ ; \*\*\* $p < 0.0001$ ; <sup>#</sup> $p > 0.05$ (Within group comparison of baseline versus week 8 data, paired t-test)
ARE: Ashwagandha, PL: Placebo, C.I.: Confidence Interval, SEM: Standard error for mean, SD: Standard deviation, FAST: Frailty index score assessment, 6MWT: Six Minute. Walking Endurance Test, MMSE: Mini-Mental State Exam, PSQI: Pittsburgh Sleep Quality Index, CK: Creatinine kinase CRP: C-Reactive protein; SF-12: Short-Form 16 quality of life questionnaire.

### Serum cortisol, CRP and creatinine kinase

The two groups were similar (p>0.05) with respect to the baseline values for serum cortisol, CRP and creatinine kinase (table-2). The serum cortisol, CRP and creatinine kinase values were similar with both ARE and placebo at week 8. However, significant reduction (p<0.0001) in these values were observed with ARE, but not placebo (p>0.05).

### Hematology and serum biochemistry

The two groups were similar (p>0.05) with respect to the baseline values for hematology (cell counts, hemoglobin, and hematocrit), and all serum biochemical assessments (table-3). No significant changes were observed with both ARE or placebo in any of the laboratory parameters.

**Table 3.**
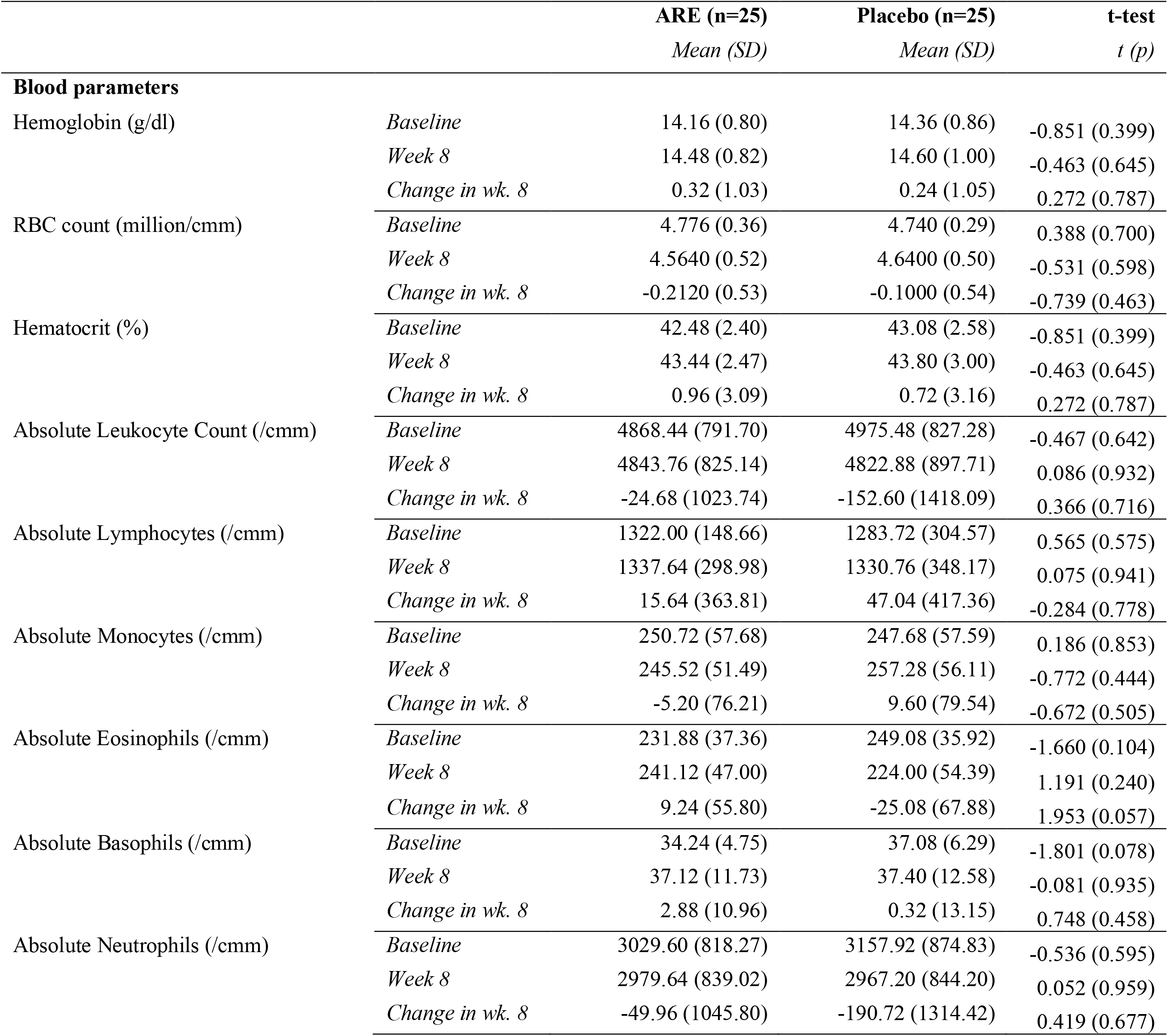
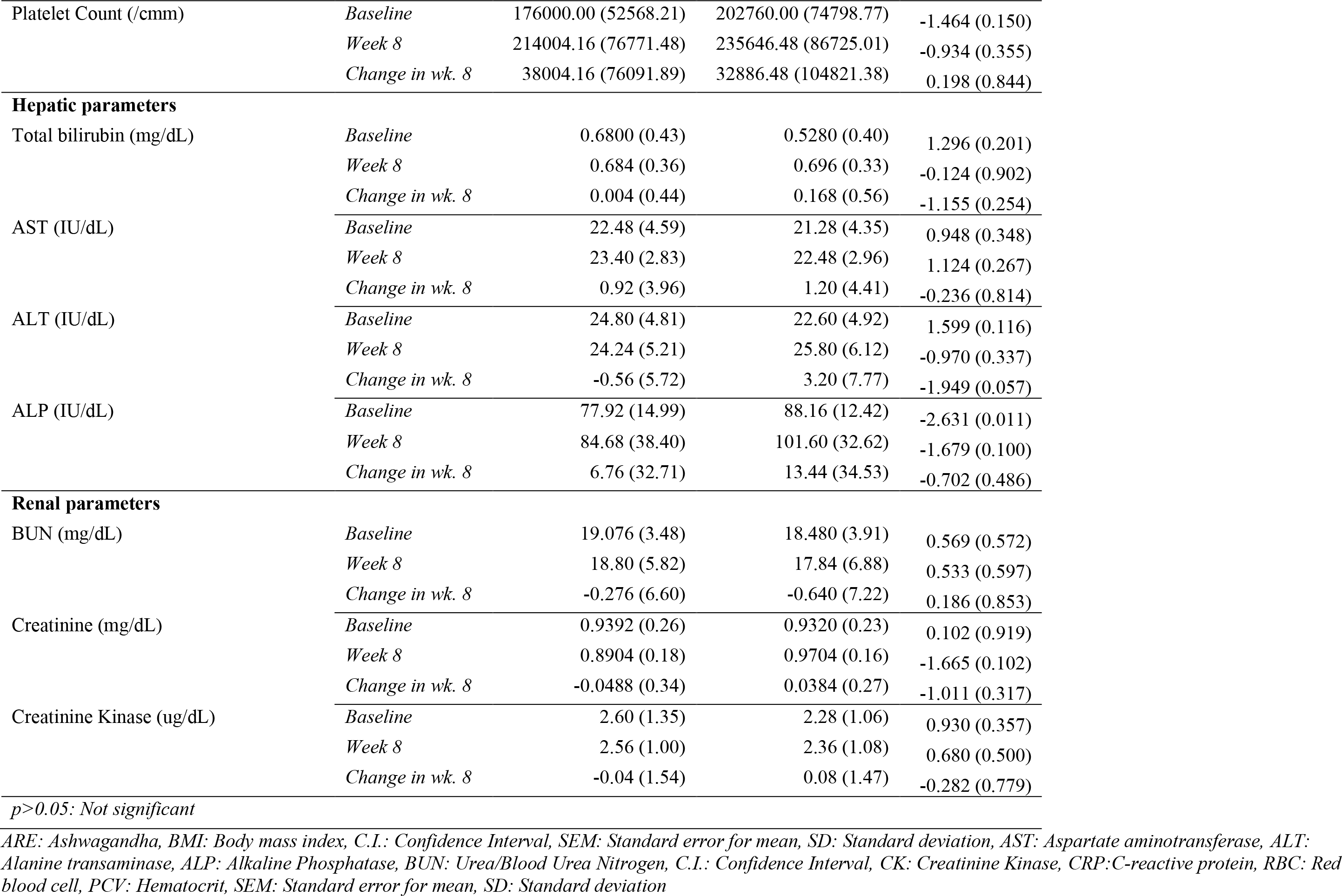
Laboratory parameters (baseline and change from baseline) in two groups.

### Adverse events

No serious adverse events (SAE) were reported during the study. However, 6 patients (5 with ARE and 1 with placebo) reported mild adverse events (AE). Patients treated with ARE reported headache (n=2), nausea (n=1), vomiting (n=1) and body pain (n=1), whereas patients in the placebo group reported stomach pain (n=1). Stomach pain was treated with oral acetaminophen, nausea was treated with domperidone, and pain was treated with analgesic and non-steroidal anti-inflammatory drugs. All the adverse events completely recovered. However, no serious adverse events/episodes were reported in the study period and during the follow-up.

## DISCUSSION

The elderly frail individuals are the main users of medical and social health care services. However, no effective therapeutic options are available to improve frailty.[18] Pharmacological options such as ACE inhibitors, testosterone, IGFs, Vitamin D, and calcium are not recommended for the treatment of frailty.[19], [20] Protein supplementation to improve muscle mass can be used in prefrail or frail malnourished elderly people, and no detrimental effect on kidney function in elderly people has been reported in one study by Park Y et al (2018).[21] However, as reported by Vasconcelos QDJS et al. (2021), long term consumption of excess protein supplementation may have some adverse effects on the body, which can further be aggravated by sedentary lifestyle.[22], [23]

Traditional Ayurveda and the modern medical literature writings report many potential health benefits of the plant Ashwagandha (*Withania somnifera*) as herb. Ashwagandha is a member of the family of herbs referred to as “adaptogens”. The term “adaptogen” is applied to an herb with phytonutrients that regulate metabolism when a body is perturbed by physical or mental stress, and help the body adapt by normalizing system functions, developing resistance to future such as stress, and elevating the body’s functioning to a higher level of performance.[24] This study assessed the efficacy of Ashwagandha root extract in the management of frailty in older adults using FAST (Frailty Assessment and Screening Tool), 6-minute walk test, MMSE, PSQI, and SF-12 scores. We observed a statistically significant reduction in FAST score (p=0.008), PSQI (p<0.001), and SF-12 score (p<0.001) with ashwagandha root extract compared with placebo. This indicates a significant reduction in stress levels. Significant improvement in sleep quality is an important indicator of better stress management. Stress management effects of Ashwagandha which helps to improve sleep quality and thus improves quality of life in older frail patients. Salve J et al (2019) have also reported significant reduction in stress as assessed on the Perceived Stress Scale (PSS scores) and serum cortisol levels with Ashwagandha 250 mg/day (p<0.05) and 600 mg/day (p<0.001). The participants who received ashwagandha showed significant improvement in sleep quality as compared to the placebo group.[25] Chandrashekhar et al (2012) also reported significant reduction in all stress assessment scale scores and serum cortisol levels. [26] Kae Ling et al documented in a systematic review and meta-analysis that ashwagandha root extract appears to have a beneficial effect in improving sleep in adults. A total of five randomized controlled trials containing 400 participants were analyzed. Ashwagandha extract exhibited a significant effect on overall sleep (Standardized Mean Difference -0.59; 95% Confidence Interval -0.75 to -0.42; I2 = 62%). It seems to be a relatively safe intervention and can be considered as a more effective option in improving sleep, with a dosage ≥600mg/d for ≥8weeks. Ashwagandha extract was also found to improve the mental alertness on rising in morning and anxiety level. [27]

The effect of ashwagandha supplementation on muscle strength and recovery has been studied in the last few years. A randomized placebo-controlled study by Wankhede S et al (2015) reported significant increases in muscle mass and strength with ashwagandha root extract supplementation. There was a significant increase in muscle strength in bench press (p=0.001) and the leg extension (p=0.04) exercises with Ashwagandha. These findings suggested that Ashwagandha supplementation may be useful in conjunction with resistant training program.[28] In our study, we assessed the aerobic capacity and physical endurance using the 6-minute walk test. We observed significant improvement in 6MWT score in ARE group (p<0.001) whereas, no change was observed with placebo (p=0.794). Further, ashwagandha cause a significant reduction in exercise induced muscle damage as indicated by the stabilization of serum creatine kinase with Ashwagandha. Also, there was greater recovery from muscle damage with Ashwagandha supplementation. This faster recovery could be attributed to the antioxidant effects to combat free radical damage both at the muscle and central nervous system levels, anti-inflammatory, analgesic and reduction in lactic acid and blood urea nitrogen caused by Ashwagandha.[29], [30], [31] Harish S et al (2021) reported significant improvement in total MMSE total and subscale scores for different domains of cognitive function with 8-week therapy with ashwagandha, suggesting a definitive role of Ashwagandha in the management of cognitive impairment and dementia.[32]

In a prospective population-based study in 1020 participants aged 65 years and older living in Italy, serum inflammatory markers CRP, IL-6, and IL1 were correlated with physical performance (r=-0.162, r=-0.251, and r=-0.127, respectively).[33] Thus, in elderly reduced physical performance is associated with increase in inflammatory markers. We observed significant reduction in serum CRP, cortisol and creatinine kinase with 8-week ashwagandha treatment. Oxidative stress and inflammation are the characteristic features of age-related muscle atrophy. Our study assessed the aerobic capacity and endurance by using 6-MWT. A significant reduction in 6MWT scores was observed with ARE group (p<0.001) whereas no reduction was observed with PL group (p=0.794). Furthermore, ARE group also had a significantly greater reduction in exercise-induced muscle damage (EIMD), stabilization of serum creatinine kinase as ARE group showed significant change 0.04 (1.54) from the baseline to week 8 as compared to PL group 0.08 (1.47) Ashwagandha has demonstrated the potential to mitigate these physiological alterations by suppressing systemic low-grade inflammation by reducing levels of IL-1β and TNFα in serum.[34] Additionally, Ashwagandha significantly increased the expression of growth factors related to muscle regeneration & mitochondrial biogenesis and thus contributes to the enhancement of muscle function, exercise endurance, muscle mass & strength. In multiple clinical trials, Ashwagandha has shown to be effective in reduction of chronic inflammation markers such as C-reactive protein (CRP).[35] This may suggest anti-inflammatory effects of Ashwagandha even in individuals with low levels of inflammation. These findings are consistent with our findings. This supports the use of Ashwagandha to prevent chronic inflammatory diseases. The decreased effect of Ashwagandha on cortisol levels may have contributed to the muscle growth and consequent increase in creatinine levels. Adrian Gomez et al (2023) reported maintenance of normal ranges for serum creatinine with Ashwagandha in their study and suggested that it was most likely due to increments in muscle mass by Withanolides present in Ashwagandha extract.[36] The increase in serum testosterone and reduced cortisol caused by Ashwagandha may have caused muscle growth with resultant increase in creatinine levels.

The adverse effects reported in this study were body-ache (4%), headache (8%), nausea (4%), vomiting (2%), and stomach pain (2%) of mild to moderate severity which were unlikely related to Ashwagandha administration and recovered completely with concomitant therapy. Kelgane SB et al (2020) also observed that Ashwagandha supplement was well tolerated and safe for the study population.[5] Several clinical studies have proven the efficacy, tolerability and safety of use of Ashwagandha in various contexts. others benefit of ARE boasts sleep quality and muscle endurance. This herbal remedy holds significant potential for elevating overall health and well-being, serving as a general tonic for the same purpose. Furthermore, when employed as a complementary treatment in various psychosomatic disorders, ashwagandha is recommended due to its capacity to enhance neuromuscular endurance, mental and physical resilience, and tissue viability. Thus, Ashwagandha looks like a potential treatment for old adults with frailty. This herbal remedy holds significant potential for elevating overall health and well-being and serving as a general tonic for the same purpose.

## STRENGTH AND LIMITATIONS

The strengths of this study include its double-blind design and >80% power of analysis of primary outcome of the study. The study cohort was narrow due to stringent eligibility criteria for participation in the study. It should be considered during interpretation of the results. Further research is needed to confirm the findings of this study in a larger & diverse group of the population.

## CONCLUSION

Ashwagandha has demonstrated an excellent efficacy & safety profile in older adults as there were no serious adverse effects & no changes in hepatic and renal parameters observed. There was a significant reduction in FAST score and improvement in, 6MWT, PSQI, MMSE & SF-12 scores at the end of the study. These findings denote the definitive role of Ashwagandha in the management of frailty-associated cognitive impairment, physical & mental vulnerability, poor sleep quality, stress, anxiety & sarcopenia. Ashwagandha showed significant potential in enhancing overall health, general well-being & quality of life in older adults. Therefore, it can be recommended as a better supplement to reduce diverse symptoms of frailty.

## Data Availability

Data will be made available based on request.

## CRediT AUTHOR STATEMENT

Author 1: Conceptualization, Investigation, Resources, Writing – Review & Editing

Author 2: Investigation, Writing – Review & Editing

Author 3: Investigation, Writing – Review & Editing

Author 4: Investigation, Writing – Review & Editing

## DATA AVAILABILITY

Data will be made available based on request.

## ACKNOWLEDGEMENTS

The authors thank Ixoreal BioMed Inc., Los Angeles, California, USA, for supplying the KSM-66 ashwagandha root extract used in the study, and Clinsearch Healthcare Solutions Pvt Ltd for manuscript writing and publication services.

## REFERENCES

[1] E. Topinková, ‘Aging, Disability and Frailty’, Ann Nutr Metab, vol. 52, no. Suppl. 1, pp. 6–11, 2008, doi: 10.1159/000115340.

[2] K. Yashoda and A. Nagarkar, ‘Prevalence and Correlates of Frailty in Older adults in India’, Indian Journal of Gerontology, vol. 30, Sep. 2016.

[3] R. B. Biritwum et al., ‘Prevalence of and factors associated with frailty and disability in older adults from China, Ghana, India, Mexico, Russia and South Africa’, Maturitas, vol. 91, pp. 8–18, 2016, doi: 10.1016/j.maturitas.2016.05.012.

[4] J. At et al., ‘Frailty and the prediction of dependence and mortality in low- and middle-income countries: a 10/66 population-based cohort study’, BMC Med, vol. 13, no. 1, p. 138, Dec. 2015, doi: 10.1186/s12916-015-0378-4.

[5] S. B. Kelgane, J. Salve, P. Sampara, and K. Debnath, ‘Efficacy and Tolerability of Ashwagandha Root Extract in the Elderly for Improvement of General Well-being and Sleep: A Prospective, Randomized, Double-blind, Placebo-controlled Study’, Cureus, Feb. 2020, doi: 10.7759/cureus.7083.

[6] S. Vermeiren et al., ‘Frailty and the Prediction of Negative Health Outcomes: A Meta-Analysis’, Journal of the American Medical Directors Association, vol. 17, no. 12, p. 1163.e1-1163.e17, 2016, doi: 10.1016/j.jamda.2016.09.010.

[7] J. T. Viitasalo, P. Era, A.-L. Leskinen, and E. Heikkinen, ‘Muscular strength profiles and anthropometry in random samples of men aged 31–35, 51–55 and 71–75 years’, Ergonomics, vol. 28, no. 11, pp. 1563–1574, Nov. 1985, doi: 10.1080/00140138508963288.

[8] A. M. Negm et al., ‘Management of frailty: a protocol of a network meta-analysis of randomized controlled trials’, Syst Rev, vol. 6, no. 1, p. 130, Dec. 2017, doi: 10.1186/s13643-017-0522-7.

[9] S. Saleem, G. Muhammad, M. A. Hussain, M. Altaf, and S. N. A. Bukhari, ‘Withania somnifera L.: Insights into the phytochemical profile, therapeutic potential, clinical trials, and future prospective’, Iranian Journal of Basic Medical Sciences, vol. 23, no. 12, Dec. 2020, doi: 10.22038/ijbms.2020.44254.10378.

[10] X. J. Zhou, D. Rakheja, X. Yu, R. Saxena, N. D. Vaziri, and F. G. Silva, ‘The aging kidney’, Kidney International, vol. 74, no. 6, pp. 710–720, Sep. 2008, doi: 10.1038/ki.2008.319.

[11] J. Salve, S. Pate, K. Debnath, and D. Langade, ‘Adaptogenic and Anxiolytic Effects of Ashwagandha Root Extract in Healthy Adults: A Double-blind, Randomized, Placebo-controlled Clinical Study’, Cureus, Dec. 2019, doi: 10.7759/cureus.6466.

[12] K. De et al., ‘Development and Psychometric Validation of a New Scale for Assessment and Screening of Frailty Among Older Indians’, CIA, vol. Volume 16, pp. 537–547, Mar. 2021, doi: 10.2147/CIA.S292969.

[13] ’ATS Statement: Guidelines for the Six-Minute Walk Test’, Am J Respir Crit Care Med, vol. 166, no. 1, pp. 111–117, Jul. 2002, doi: 10.1164/ajrccm.166.1.at1102.

[14] D. J. Buysse, C. F. Reynolds, T. H. Monk, S. R. Berman, and D. J. Kupfer, ‘The Pittsburgh sleep quality index: A new instrument for psychiatric practice and research’, Psychiatry Research, vol. 28, no. 2, pp. 193–213, 1989, doi: 10.1016/0165-1781(89)90047-4.

[15] K. Sallam, ‘The Use of the Mini-Mental State Examination and the Clock-Drawing Test for Dementia in a Tertiary Hospital’, JCDR, 2013, doi: 10.7860/JCDR/2013/4203.2803.

[16] M. F. Folstein, S. E. Folstein, and P. R. McHugh, ‘“Mini-mental state”‘, Journal of Psychiatric Research, vol. 12, no. 3, pp. 189–198, Nov. 1975, doi: 10.1016/0022-3956(75)90026-6.

[17] D. Turner-Bowker and S. J. Hogue, ‘Short Form 12 Health Survey (SF-12)’, in Encyclopedia of Quality of Life and Well-Being Research, A. C. Michalos, Ed., Dordrecht: Springer Netherlands, 2014, pp. 5954–5957. doi: 10.1007/978-94-007-0753-5_2698.

[18] G. Kojima, A. Liljas, and S. Iliffe, ‘Frailty syndrome: implications and challenges for health care policy’, RMHP, vol. Volume 12, pp. 23–30, Feb. 2019, doi: 10.2147/RMHP.S168750.

[19] E. Dent et al., ‘Physical Frailty: ICFSR International Clinical Practice Guidelines for Identification and Management’, The Journal of nutrition, health and aging, vol. 23, no. 9, pp. 771–787, Nov. 2019, doi: 10.1007/s12603-019-1273-z.

[20] A. Clegg, J. Young, S. Iliffe, M. O. Rikkert, and K. Rockwood, ‘Frailty in elderly people’, The Lancet, vol. 381, no. 9868, pp. 752–762, Mar. 2013, doi: 10.1016/S0140-6736(12)62167-9.

[21] Y. Park, J.-E. Choi, and H.-S. Hwang, ‘Protein supplementation improves muscle mass and physical performance in undernourished prefrail and frail elderly subjects: a randomized, double-blind, placebo-controlled trial’, The American Journal of Clinical Nutrition, vol. 108, no. 5, pp. 1026– 1033, Nov. 2018, doi: 10.1093/ajcn/nqy214.

[22] K. Sukalingam, K. Ganesan, and S. Das, ‘An insight into the harmful effects of soy protein: A review’, La Clinica Terapeutica, no. 3, pp. 131–139, Jun. 2015, doi: 10.7417/CT.2015.1843.

[23] Q. D. J. S. Vasconcelos, T. P. R. Bachur, and G. F. Aragão, ‘Whey protein supplementation and its potentially adverse effects on health: a systematic review’, Appl. Physiol. Nutr. Metab., vol. 46, no. 1, pp. 27–33, Jan. 2021, doi: 10.1139/apnm-2020-0370.

[24] K. Abascal and E. Yarnell, ‘Increasing Vitality with Adaptogens: Multifaceted Herbs for Treating Physical and Mental Stress’, Alternative and Complementary Therapies, vol. 9, no. 2, pp. 54–60, Apr. 2003, doi: 10.1089/107628003321536959.

[25] J. Salve, S. Pate, K. Debnath, and D. Langade, ‘Adaptogenic and Anxiolytic Effects of Ashwagandha Root Extract in Healthy Adults: A Double-blind, Randomized, Placebo-controlled Clinical Study’, Cureus, Dec. 2019, doi: 10.7759/cureus.6466.

[26] K. Chandrasekhar, J. Kapoor, and S. Anishetty, ‘A Prospective, Randomized Double-Blind, Placebo-Controlled Study of Safety and Efficacy of a High-Concentration Full-Spectrum Extract of Ashwagandha Root in Reducing Stress and Anxiety in Adults’, Indian Journal of Psychological Medicine, vol. 34, no. 3, pp. 255–262, Jul. 2012, doi: 10.4103/0253-7176.106022.

[27] K. L. Cheah, M. N. Norhayati, L. Husniati Yaacob, and R. Abdul Rahman, ‘Effect of Ashwagandha (Withania somnifera) extract on sleep: A systematic review and meta-analysis’, PLoS ONE, vol. 16, no. 9, p. e0257843, Sep. 2021, doi: 10.1371/journal.pone.0257843.

[28] S. Wankhede, D. Langade, K. Joshi, S. R. Sinha, and S. Bhattacharyya, ‘Examining the effect of Withania somnifera supplementation on muscle strength and recovery: a randomized controlled trial’, Journal of the International Society of Sports Nutrition, vol. 12, no. 1, p. 43, 2015.

[29] L. C. Mishra, B. B. Singh, and S. Dagenais, ‘Scientific basis for the therapeutic use of Withania somnifera (ashwagandha): a review’, Altern Med Rev, vol. 5, no. 4, pp. 334–346, Aug. 2000.

[30] V. Vyas, P. Bhandari, and R. Patidar, ‘A Comprehensive Review on Withania somnifera Dunal’, Journal of Natural Remedies, vol. 11, pp. 1–13, Jan. 2011, doi: 10.18311/jnr/2011/43.

[31] G. Singh, P. K. Sharma, R. Dudhe, and S. Singh, ‘Biological activities of Withania somnifera’, Ann Biol Res, vol. 1, Nov. 2009.

[32] Dr. H. S. * Dr. Harish S.*, Madhu Krishnamani Madhu Krishnamani, ‘A CLINICAL STUDY TO EVALUATE COMPARATIVE CLINICAL EFFICACY AND SAFETY OF BHC9612CP ON COGNITIVE FUNCTION IN ADULT SUBJECTS’, EJPMR, vol. ejpmr, 2021,8(7), 404–414, Jun. 2021.

[33] M. Cesari et al., ‘Inflammatory Markers and Physical Performance in Older Persons: The InCHIANTI Study’, The Journals of Gerontology Series A: Biological Sciences and Medical Sciences, vol. 59, no. 3, pp. M242–M248, Mar. 2004, doi: 10.1093/gerona/59.3.M242.

[34] E. Logie and W. Vanden Berghe, ‘Tackling Chronic Inflammation with Withanolide Phytochemicals—A Withaferin A Perspective’, Antioxidants, vol. 9, no. 11, p. 1107, Nov. 2020, doi: 10.3390/antiox9111107.

[35] B. Auddy, J. Hazra, A. Mitra, B. Abedon, and S. Ghosal, ‘A Standardized Withania Somnifera Extract Significantly Reduces Stress-Related Parameters in Chronically Stressed Humans: A Double-Blind, Randomized, Placebo-Controlled Study’, Journal of American Nutraceutical Association, vol. 11, pp. 50–56, Nov. 2008.

[36] A. Gómez Afonso, D. Fernandez-Lazaro, D. P. Adams, A. Monserdà-Vilaró, and C. I. Fernandez-Lazaro, ‘Effects of Withania somnifera (Ashwagandha) on Hematological and Biochemical Markers, Hormonal Behavior, and Oxidant Response in Healthy Adults: A Systematic Review’, Curr Nutr Rep, vol. 12, no. 3, pp. 465–477, Jul. 2023, doi: 10.1007/s13668-023-00481-0.

